# Entity-centric evaluation of large language model responses for medical question-answering tasks

**DOI:** 10.1101/2025.11.12.25340106

**Authors:** Yi Liu, Vijaya B. Kolachalama

**Affiliations:** Faculty of Computing & Data Sciences, Boston University, MA, USA; Department of Medicine, Boston University Chobanian & Avedisian School of Medicine, Boston, MA, USA; Department of Computer Science, Boston University, MA, USA

## Abstract

**Objective:** Develop a metric for evaluating the clinical alignment and informativeness of large language model (LLM)-generated responses in medical question-answering (QA) tasks.

**Materials and methods:** We propose EntQA, an entity-centric metric that extracts biomedical entities from patient backgrounds, diagnostic questions and LLM responses using a biomedical named entity recognition model, followed by de-duplication and semantic/lexical matching with thresholds. We computed recall-style coverage scores to quantify entity retention and detect omissions without external resources. We evaluated EntQA on five benchmarks using seven Qwen 2.5 Instruct models (0.5B–72B parameters), comparing it to baselines via Spearman/Kendall correlations with model accuracy at group level, point-biserial correlations at case level, and Spearman correlations with model scaling.

**Results:** EntQA demonstrated consistent positive alignments with accuracy (group-level Spearman up to 0.9286; case-level point-biserial up to 0.0926) and model scaling (Spearman up to 0.252), outperforming baselines which often showed negative or inconsistent correlations (e.g., BERTScore Spearman -0.9286 with accuracy).

**Conclusion:** EntQA offers a scalable, interpretable evaluation for LLM medical QA, outperforming traditional metrics in capturing clinical fidelity and supporting trustworthy healthcare AI through applications in fact-checking and model refinement.

## 1 Introduction

Large language models (LLMs) have demonstrated strong performance on a range of medical question-answering (QA) tasks, including those that emulate licensing exams such as the USMLE. For instance, models like Med-PaLM^1^ have achieved expert-level accuracy by exceeding passing thresholds on USMLE-style questions, showcasing their potential to assist in diagnostic reasoning and knowledge retrieval. However, existing evaluation approaches often rely on coarse metrics such as accuracy or BLEU scores^2^, which offer limited insight into the clinical relevance or informativeness of model-generated responses. These metrics do not capture aspects such as the depth of rationale or adherence to patient-specific details, which are critical for practical deployment. In real-world clinical settings, the quality of a response is not solely judged by whether the final answer is correct, but also by how well the rationale reflects key patient-specific factors, such as symptoms, medical history, and contextual evidence, alongside sound medical reasoning. LLM outputs may omit critical clinical details (e.g., overlooking family history or comorbidities), introduce hallucinations (e.g., fabricating non-existent test results), or rely on generic reasoning that deviates from the input context (e.g., providing broad overviews instead of tailored explanations), potentially misleading users, contributing to diagnostic errors, and limiting the trustworthiness of such systems in high-stakes environments such as medicine.

Prior work on evaluating LLMs in medical question answering has predominantly focused on benchmarks targeting multiple-choice formats, such as MedQA^3^ and USMLE-style tasks^4^, where accuracy was the primary evaluation metric. Later work expanded the evaluation scope and evaluation metrics, moving beyond static, closed-ended questions to more realistic, open-ended clinical tasks that require reasoning, summarization, and contextual understanding. Benchmarks such as MedBench^5^ and Med-S-Bench^6^ broaden task coverage, ranging from information extraction, closed-ended diagnosis to open-ended summarization, treatment explanation and report generation. These aspects reflect the complexity of real-world clinical applications, while MedExpQA^7^ and MMedBench^8^ additionally evaluate LLMs in multilingual settings. Additionally, metrics evolved to evaluate free-form text, adopting string-overlap metrics (e.g., BLEU^2^, ROUGE^9^, METEOR^10^) and embedding-based measures (e.g., BERTScore^11^) to capture semantic similarity between LLM’s response with ground-truth references. While high-quality, expert written references are ideal, they are often costly to obtain on scale. As a result, many studies use existing clinical texts as proxies. For example, some studies used the conclusion section of research articles or the impression section of radiology reports as ground-truth summaries in medical summarization tasks^6^. In order to utilize the existing clinical standard to measure the usefulness of LLM response, some studies^12^ introduced detailed rubrics such as factual accuracy, completeness, clarity, contextual awareness, and instruction adherence, sometimes involving physician raters^13,14^, and other times relying on advanced LLM graders^12^. In addition, other studies have emphasized specific evaluation perspectives such as hallucination detection and logical validity. For hallucination detection, some studies decompose LLM’s generated outputs into verifiable units, either by extracting key medical entities^15^ or decomposing multi-step reasoning chains^14,16,17,^ and assess the factuality of each component using external sources, either a local literature database (e.g., PubMed), online search or some structured knowledge graph ^18^(e.g. UMLS graph). To assess reasoning validity, studies tracked whether new information is introduced across generation steps and used uncertainty-based metrics^19^ or agentic classifiers ^16^ to detect contradictions or unnecessary repetition relative to prior reasoning steps. Despite these advancements, current approaches often depend on external databases or auxiliary models, limiting scalability and interpretability of how response quality is judged. More importantly, they lack a fine-grained, interpretable quantification of how well an LLM’s reasoning aligns with patient context and diagnostic intent, which is central to clinical decision support.

Here, we introduce EntQA, an entity-centric metric tailored for medical QA tasks that involve patient background information and a diagnostic inquiry. Unlike existing approaches that rely on external databases or auxiliary models, EntQA operates solely on the existing task context including the question, background, and LLM-generated responses, using biomedical named entity recognition to extract clinically relevant entities (e.g., symptoms, diagnoses, treatments) and query’s main intent. It then computes recall-style coverage from two perspectives: (1) EntQA_background_ or EntQA_b_ quantifies how well the response incorporates relevant contextual information from the patient’s clinical history, and (2) EntQA_question_ or EntQA_q_ measures how effectively it addresses the diagnostic focus of the question. These recall scores provide interpretable insights into whether the LLM’s reasoning is adequately grounded in the patient’s context and the clinical inquiry, as the pipeline further alerts which key clinical entities are missing from the response, ensuring that responses remain trustworthy and free from topical drift. To foster proper evaluation, we curated a dataset of LLM-generated responses for USMLE-style questions. We performed a meta-evaluation of EntQA against other metrics using this dataset and additional benchmarks, where it consistently showed higher correlation with model accuracy and reasoning ability.

## 2 Methods

We propose **EntQA**, an entity-centric framework tailored for medical benchmark QA tasks, which quantifies how effectively large language models (LLMs) interpret and incorporate clinically relevant information from input to their generated summaries. Unlike generic n-gram overlap metrics, EntQA explicitly measures the proportion of key medical entities - derived from either patient background or diagnostic question - that are preserved in the model’s response. This allows us to compare different LLM-generated summaries with respect to medical coverage and clinical reasoning fidelity.

As illustrated in Figure 1, the EntQA pipeline consists of three stages: (1) entity extraction and refinement, (2) entity sematic matching, and (3) coverage score computation. Importantly, because medical benchmark inputs are long and contain both patient background (B) and a real diagnostic question (Q), we design EntQA from two complementary perspectives to evaluate LLM’s response (A): EntQA using background as input and EntQA using question as input.

**Figure 1:**
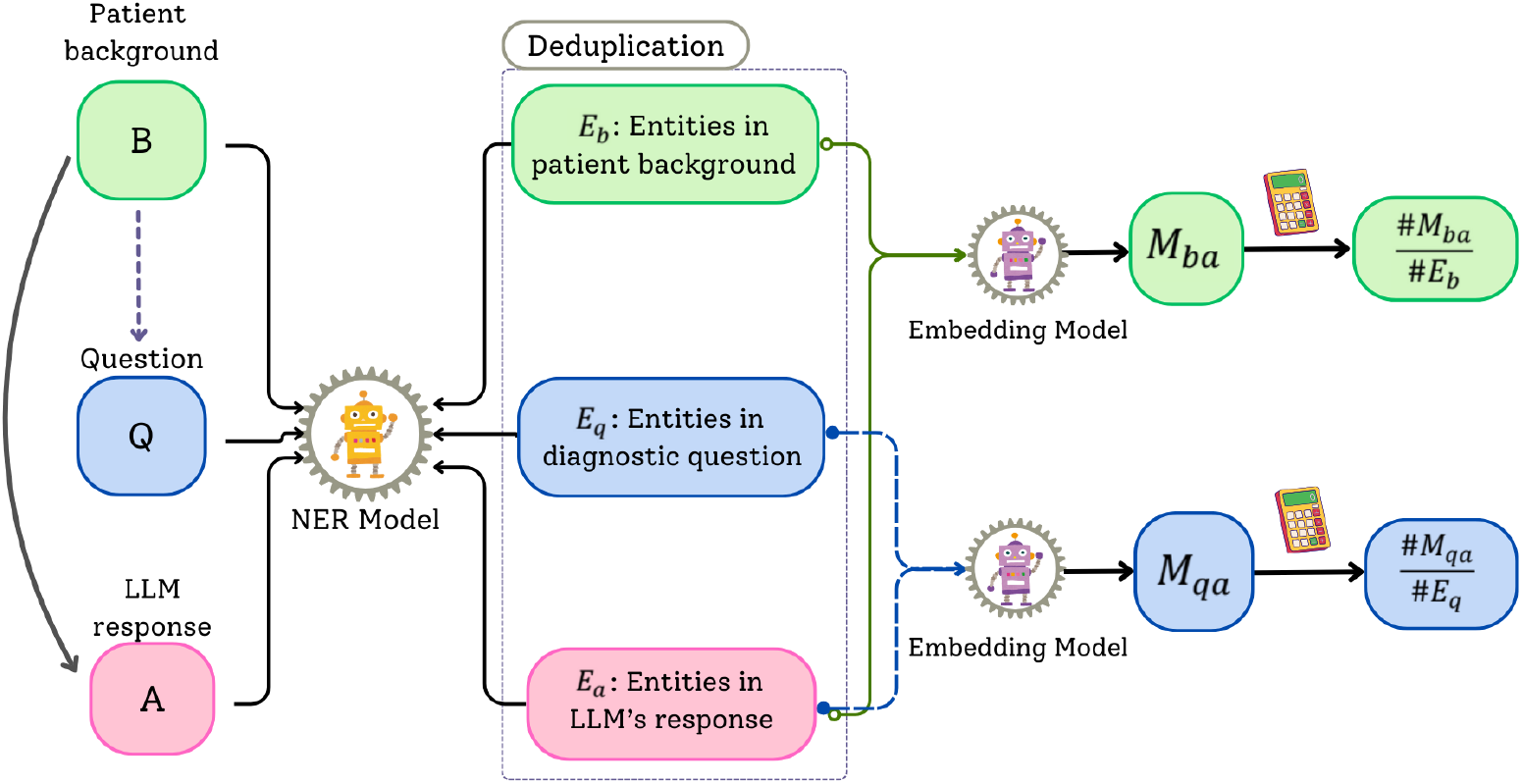
Schematic of the entity-based evaluation pipeline for LLM-generated responses. Named entity recognition (NER) extracts entities from the patient background (*E*_*b*_), diagnostic question (*E*_*q*_), and LLM response (*E*_*a*_). Entities from all three sources undergo de-duplication before embedding. An embedding model computes semantic matches, yielding coverage metrics: the fraction of matched background entities in the response 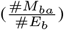 and matched question entities in the response 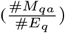.

1. <u>Structured entity extraction.</u> We begin by separating the benchmark input into two components: the patient background (B) and the diagnostic question (Q). From these, as well as the LLM-generated answer (A), we extracted clinically relevant entities using a biomedical NER model^20^. This NER model captures a wide range of medical concepts, including demographic information, clinical events, diagnostic procedures, disease disorders, and biological structures. This step yields three sets of entities:
  - *E*_*b*_: entities extracted from the patient background,
  - *E*_*q*_: entities extracted from the diagnostic question,
  - *E*_*a*_: entities extracted from the LLM-generated answer. We further refine the noise and redundancy in the raw entities by removing entities whose surface forms are equivalent up to punctuation or minor orthographic variation by retaining the longest mention; and filter out entities that are vague, repetitive, or generic and introduced by multiple-choice question stem.
2. <u>Entity semantic matching.</u> Next, we determine whether the input entities are faithfully represented in the LLM summary. We employ a state-of-the-art embedding model to compute semantic similarity between entity pairs. Two matchings are performed:
  - *M*_*ba*_: matches between entities in patient background *E*_*b*_ and answer entities *E*_*a*_,
  - *M*_*qa*_: matches between entities in diagnostic question *E*_*q*_ and answer entities *E*_*a*_, An entity pair is considered matched if (i) their embedding similarity exceeds a threshold (e.g., cosine similarity *>* 0.7), and (ii) they share sufficient lexical overlap (e.g., token-level overlap *>* 30%).
3. <u>Coverage score computation.</u> Finally, we compute the EntQA coverage score as a recall-style measure:
  - Background perspective:

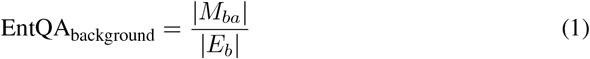

i.e., the proportion of patient background entities captured and represented in the LLM summary.
  - Question perspective:

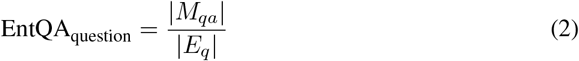

i.e., the proportion of diagnostic question entities captured and represented in the LLM summary.

This score ranges from 0 to 1, where higher values indicate a greater degree of information interpretation and utilization. Importantly, the metric does not reward hallucinations or irrelevant details—it only quantifies how well the model retains and reflects essential source concepts.

By reporting both perspectives, EntQA not only assesses whether an LLM-generated summary adequately incorporates *contextual patient information* and addresses the *explicit diagnostic question*, but also enables users to explicitly trace which clinically important entities are preserved or omitted. This traceability provides additional information on the reliability of the summary, making EntQA a more clinically grounded and interpretable metric for benchmarking LLMs in medical QA tasks.

### Experimental settings

To systematically evaluate the reliability of large language models as AI-assisted diagnostic tools, we employed seven models from Qwen 2.5 Instruct family, which covers a wide range of parameter sizes from 0.5B to 72B. By deliberately covering this wide spectrum of parameter scales, we obtained model responses that reflect a gradient of reasoning abilities and answer qualities—ranging from shallow, error-prone rationales produced by small models to more coherent, clinically informed explanations from large models. This spectrum of outputs is critical for our study: it allows us to rigorously test whether the proposed metrics can reliably distinguish between high- and low-quality summaries by assessing how comprehensively patient-related entities are captured and reflected in the output as model capacity increases.

We collected LLM-generated response over five medical QA benchmarks that are widely recognized in medical AI research: USMLE Step 1, USMLE Step 2, USMLE Step 3^4^, MedQA^3^ and MedExpQA^7^. Each benchmark includes clinical case descriptions, multiple choice options, and the correct answers. Collectively, they form a clinically validated spectrum of test cases, covering preclinical sciences, case-based diagnostic reasoning, and advanced decision-making in patient care. The USMLE is a three-step licensing examination for medical practice in the United States. Step 1 tests core biomedical sciences such as physiology, pharmacology, and pathology (94 cases). Step 2 assesses clinical reasoning and patient management under supervision (109 cases). Step 3 evaluates advanced decision-making for independent practice, including diagnosis, prognosis, and treatment (122 cases). Together, these steps form a progressive testbed of medical competencies. MedQA is a curated benchmark collected after the USMLE board exams. The English subset we used contains 1,273 multiple-choice questions, aiming to assess a physician’s ability to apply medical knowledge, concepts and apply fundamental patient-centered skills. MedExpQA is derived from the Spanish national residency medical exam and contains 125 test cases with five answer choices and detailed explanations.

To ensure consistency of the generated output format across benchmarks, we standardized the prompt to elicit structured reasoning. Each question (patient information, question, and options) was presented to the model, followed by explicit instructions:

~~~
{background}{option}
Select the best answer and respond with:
“The correct answer is [OPTION LETTER].”
Then, write “Explanation:” and provide a detailed explanation,
clearly stating why each option is correct or incorrect.
~~~

This prompt ensures that responses provide both a definitive answer and a comprehensive explanation, allowing a downstream evaluation of the coverage, informativeness, and diagnostic validity of the entity. For all LLM models, response generation was performed under a deterministic decoding configuration with temperature = 0, top_p = 1, and maximum output length set to 1024 tokens. These parameters minimize randomness in outputs and guarantee comparability across models and benchmarks.

To systematically evaluate whether our proposed metric can reliably assess the relative quality of multiple LLM responses to the same medical question, we designed a series of experiments from two perspectives: (i) alignment with answer accuracy and (ii) alignment with model size as a proxy for LLM’s diagnostic reasoning ability. Rather than focusing solely on correlation strength, we used these analyzes to investigate whether our metric identifies summaries with greater clinical value - that is, summaries that reflect patient information more comprehensively and are more aligned with accurate diagnostic reasoning. Furthermore, we benchmarked our metrics against widely used n-gram overlap metrics (BLEU^2^, ROUGE^**?**^, METEOR^10^) to assess whether an entity-centric coverage approach better captures both the comprehensive interpretation of contextual information and the extent to which the model’s reasoning process incorporates the content of the question.

To assess whether the proposed metrics align with model correctness, we performed two complementary analyses: (1.) Group-level correlation. For each benchmark, we analyzed summaries generated by seven Qwen-2.5 Instruct models of increasing sizes. We computed the model-level accuracy and the corresponding average values of all metrics, including our proposed entity-centric metrics on background coverage (*EntQA*_*background*_ or *EntQA*_*b*_) and question (*EntQA*_*question*_ or *EntQA*_*q*_) coverage and seven baseline metrics (BLEU, ROUGE, METEOR, BERTScore^11^) and then calculated Spearman and Kendall correlation coefficients between accuracy and each metric. This analysis assesses whether models with higher diagnostic accuracy tend to produce summaries that receive higher scores from each metric, thereby indicating alignment between metric evaluations and overall model performance. (2.) Case-level correlation. To further evaluate the discriminative ability of the metrics, we aggregated predictions from all models (regardless of size) and examined the association between the binary accuracy label (correct/incorrect) and each metric score for every test case. Specifically, we applied point-biserial correlation to quantify whether higher metric values are associated with correct predictions. This analysis effectively tests whether the metrics assign significantly higher scores to true predictions than to false ones, thereby capturing their ability to separate correct from incorrect responses at the individual case level.

Beyond correctness, we examined whether the metrics can serve as indicators of response quality improvements that arise with model scaling. Specifically, we grouped the Qwen-2.5 Instruct models into four parameter categories (*<* 3B, ∼ 10B, 32B, and 72B), computed the average metric scores across all benchmarks, and then measured the Spearman correlation between model size and average metrics scores within the group across all benchmarks to assess to alignment between model scaling and metric behavior. This analysis evaluates whether larger models, with stronger reasoning capacity, are consistently rewarded with higher scores by the metrics.

## 3 Results

To illustrate the practical utility of EntQA in identifying clinically relevant overlaps and gaps in LLM-generated responses, we present a qualitative case study from MedQA (Figure 2). The scenario describes a 15-year-old boy evaluated for learning disability, poor academic performance, and social interaction difficulties, with a history of hyperactivity, poor scores on reading and writing assessments, and molecular analysis showing increased CGG trinucleotide repeats. Physical examination findings include a long face and large everted ears, almond-shaped eyes with downslanted palpebral fissures, a thin upper lip and receding chin, among others. The diagnostic question pertains to the most likely physical examination finding in this patient, with options: (A) Frontal balding and cataracts, (B) Long face and large everted ears, (C) Almond-shaped eyes and downturned mouth, (D) Thin upper lip and receding chin. The model’s response suggests Fragile X Syndrome based on symptoms like intellectual disability, social interaction issues, and physical characteristics such as long face and large ears. However, entity matching reveals covered entities (e.g., “learning disability,” “hyperactivity,” “poor scores,” “molecular analysis,” “CGG trinucleotide repeats”) and uncovered entities (e.g., “physical examination,” “teachers,” “poor scores,” “assessments” from the background; “physical examination” from the question). This yields coverage metrics of EntQA_b_ = 0.62 (fraction of matched background entities in the answer) and EntQA_q_ = 0.67 (fraction of matched question entities in the answer), highlighting that the model’s summary misses key details like the long face and large everted ears, confirming option B as the correct answer.

**Figure 2:**
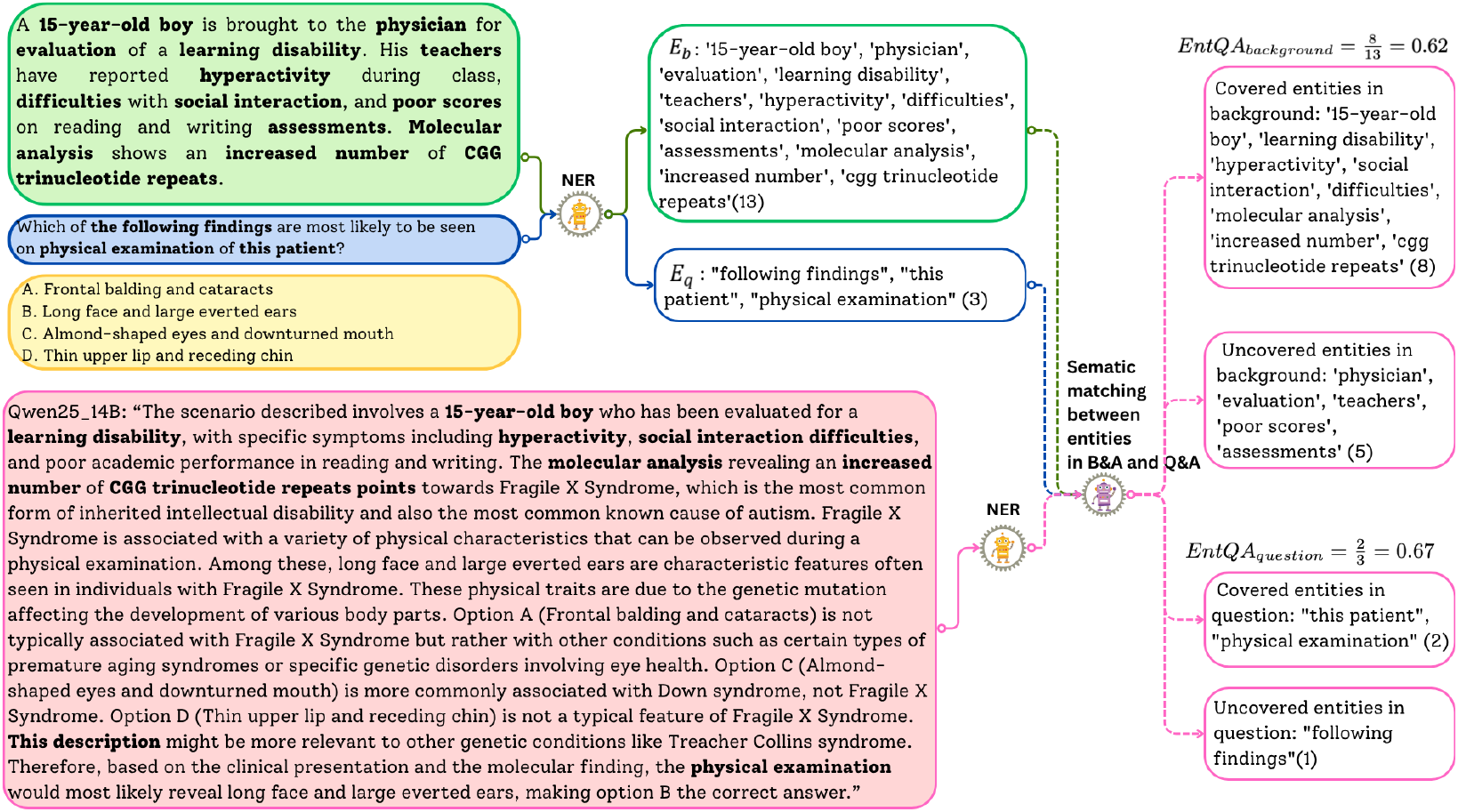
Case study from MedQA. This case demonstrates the entity-centric coverage metric’s ability to identify clinically relevant overlaps and gaps between the patient background, diagnostic question, and model-generated summary using biomedical named entity recognition (NER) and embedding-based entity matching.

### Metric alignment with accuracy

At the group level, our proposed entity-centric metrics exhibit consistent and statistically significant alignment with model correctness across all five medical QA benchmarks (Table 1). Specifically, EntQA_b_ shows consistently strong and significant positive correlations with accuracy: on MedQA, it achieves Spearman = 0.8571 (*p <* 0.05) and Kendall = 0.7143 (*p <* 0.05); on MedExpQA, it reaches the highest values among all metrics (Spearman = 0.9286, Kendall = 0.8095, *p <* 0.01); and it remains strongly positive on USMLE Step 1 (0.8571 / 0.7143) and Step 3 (0.9286 / 0.8095). Even on Step 2, where most baseline metrics degrade, EntQA_b_ still maintains a positive trend (0.7143 / 0.619). The EntQA_q_ metric also maintains positive trends(e.g., 0.6429 on MedQA, 0.7143 on Step 3), suggesting that it effectively captures how well the generated answers align with the key entities in the question and is strengthened as the model’s final prediction accuracy improves. By contrast, all baseline metrics, i.e., BLEU, ROUGE, METEOR, and BERTScore, show predominantly negative correlation with accuracy, often at statistically significant levels (e.g., ROUGE-1 get -0.8929 correlation on MedQA; METEOR’s correlation is -0.9286 on USMLE STEP 1; BERTScore’s correlation is -0.9286 on STEP 1 and -0.8571 on STEP 3). These results highlight that conventional surface-matching metrics often misalign with accuracy, whereas our entity-centric metrics provide a more faithful reflection of model reliability.

**Table 1:** Correlation between model-level accuracy and average metric scores across medical benchmarks. This table presents Spearman and Kendall correlation coefficients between model-level accuracy and average scores for various metrics on datasets including MedQA, MedExpQA, USMLE Step 1, USMLE Step 2, and USMLE Step 3. Metrics evaluated include entity-based coverage (EntQA_b_ and EntQA_q_), lexical overlap measures (BLEU-1, BLEU-4, ROUGE-1, ROUGE-2, ROUGE-L, METEOR), and semantic similarity (BERTScore-F1). Significance levels are indicated by * (*p <* 0.05) and ** (*p <* 0.01), highlighting the strength and direction of associations, with positive correlations for entity-centric metrics and often negative for traditional *n*-gram based metrics.

| Metric | MedQA |  | MedExpQA |  | USMLE_Step_1 |  | USMLE_Step_2 |  | USMLE_Step_3 |  |
| --- | --- | --- | --- | --- | --- | --- | --- | --- | --- | --- |
|  | Spearman | Kendall | Spearman | Kendall | Spearman | Kendall | Spearman | Kendall | Spearman | Kendall |
| EntQA<sub>b</sub> | 0.8571* | 0.7143* | 0.9286** | 0.8095* | 0.8571* | 0.7143 | 0.7143 | 0.619 | 0.9286** | 0.8095* |
| EntQA<sub>q</sub> | 0.6429 | 0.4286 | 0.5357 | 0.4286 | 0.6429 | 0.5238 | 0.2143 | 0.1429 | 0.7143 | 0.5238 |
| BLEU-1 | -0.8571* | -0.7143* | -0.7143 | -0.619 | -0.8214* | -0.7143* | -0.8571* | 0.7143* | 0.6429 | -0.4286 |
| BLEU-4 | 0.4643 | 0.3333 | -0.25 | -0.1429 | -0.7857* | -0.619 | -0.4286 | -0.3333 | -0.1786 | -0.1429 |
| ROUGE-1 | -0.8929** | -0.8095* | -0.8571* | -0.7143* | -0.8214* | -0.7143* | -0.8929** | -0.8095* | -0.6786 | -0.5238 |
| ROUGE-2 | -0.2857 | -0.2381 | -0.3929 | -0.3333* | -0.9286** | -0.8095* | -0.8929** | -0.7143* | -0.75 | -0.5238 |
| ROUGE-L | -0.8214* | -0.6190 | -0.6429 | -0.5238 | -0.7857* | -0.619 | -0.8571* | -0.7143* | -0.6429 | -0.4286 |
| METEOR | -0.8571* | -0.7143* | -0.6429 | -0.5238 | -0.9286** | -0.8095* | -1** | -1** | -0.75 | -0.5238 |
| BERTScore-F1 | -0.8929** | -0.8095* | -0.8929** | -0.8095* | -0.9286** | -0.8095* | -0.8929** | -0.8095* | -0.8571* | -0.7143* |

To further validate these findings beyond the aggregated model groups, Table 2 shows that at the per-case level, our two *EntQA* metrics generally achieve stronger correlations with accuracy than the baseline metrics and remain consistently positive across the benchmarks, while the conventional baseline metrics are predominantly negative. EntQA_b_ shows positive correlations on all five datasets, reaching statistical significance on MedQA (*r* = 0.0595, *p <* 0.01), MedExpQA (*r* = 0.0926, *p <* 0.01) and USMLE Step 3 (*r* = 0.0490, *p* = 0.0085). Likewise, EntQA_q_ remains positive across benchmarks and achieves significance on both MedQA (*r* = 0.0373, *p <* 0.01) and USMLE Step 3 (*r* = 0.0729, *p <* 0.05). By contrast, ROUGE-1, ROUGE-2, ROUGE-L, and BERTScore-F1 all show significant negative correlations across multiple benchmarks. (e.g., ROUGE-1 on Step 1: *r* = −0.1105, *p <* 0.01; BERTScore-F1 on Step 1: *r* = −0.1287, *p <* 0.01), and the only baseline with a positive coefficient, BLEU-4, does so on MedExpQA (*r* = 0.0987, *p <* 0.01) but fails to maintain this across other benchmarks. Taken together, these findings demonstrate that, at the case level, our EntQA metrics exhibit more robust and directionally consistent associations with case-level correctness than traditional overlap-based metrics, which frequently inversely correlated with true answer accuracy.

**Table 2:** Point-biserial correlation between per-example accuracy labels and metric scores across medical benchmarks. This table presents point-biserial correlation coefficients (*r*) and their corresponding *p*-values between per-example accuracy labels and scores for various metrics on datasets including MedQA, MedExpQA, USMLE Step 1, USMLE Step 2, and USMLE Step 3. Metrics evaluated include entity-based coverage (EntQA_b_ and EntQA_q_), lexical overlap measures (BLEU-1, BLEU-4, ROUGE-1, ROUGE-2, ROUGE-L, METEOR), and semantic similarity (BERTScore F1). Significance levels are indicated by * (*p <* 0.05) and ** (*p <* 0.01), showing generally positive correlations for entity-centric metrics and mixed or negative associations for traditional *n*-gram based metrics.

| Metric | MedQA |  | MedExpQA |  | USMLE_Step_1 |  | USMLE_Step_2 |  | USMLE_Step_3 |  |
| --- | --- | --- | --- | --- | --- | --- | --- | --- | --- | --- |
| | $r$ | $p$ | $r$ | $p$ | $r$ | $p$ | $r$ | $p$ | $r$ | $p$ |
| EntQA<sub>b</sub> | 0.0595** | 0.0 | 0.0926** | 0.0061 | 0.0129 | 0.7425 | 0.0394 | 0.2769 | 0.09** | 0.0085 |
| EntQA<sub>q</sub> | 0.0373** | 0.0004 | 0.0222 | 0.5113 | 0.0055 | 0.889 | -0.0236 | 0.5159 | 0.0729* | 0.0332 |
| BLEU-1 | -0.0593** | 0.0 | -0.0068 | 0.8410 | -0.0977* | 0.0123 | -0.1153** | 0.0014 | 0.0071 | 0.8367 |
| BLEU-4 | 0.0113 | 0.2866 | 0.0987** | 0.0035 | -0.0389 | 0.3196 | -0.0327 | 0.3676 | 0.0081 | 0.8140 |
| ROUGE-1 | -0.0667** | 0.0 | -0.0050 | 0.8828 | -0.1105** | 0.0046 | -0.1517** | 0.0000 | -0.0113 | 0.7408 |
| ROUGE-2 | -0.0028 | 0.7905 | 0.0532 | 0.1156 | -0.0596 | 0.1274 | -0.1092** | 0.0025 | -0.0011 | 0.9746 |
| ROUGE-L | -0.0425** | 0.0001 | 0.0206 | 0.5419 | -0.1050** | 0.0071 | -0.1434** | 0.0001 | -0.0171 | 0.6183 |
| METEOR | -0.0242* | 0.0225 | 0.0273 | 0.4201 | -0.0533 | 0.1724 | -0.1373** | 0.0001 | -0.0005 | 0.9875 |
| BERTScore_F1 | -0.0826** | 0.0 | -0.0435 | 0.1991 | -0.1287** | 0.0010 | -0.1018** | 0.0049 | -0.0639 | 0.0622 |

### Metrics under model scaling

Our proposed entity-centric metrics exhibit consistent positive alignment with model scaling across all five medical QA benchmarks (Table 3). EntQA_b_ shows significant positive correlations with model size on every dataset, including 0.188 on MedQA, 0.181 on MedExpQA, 0.123 on Step 1, 0.171 on Step 2, and 0.252 on Step 3, demonstrating that background-level entity coverage systematically improves as models become larger and more capable. EntQA_q_ also trends positive, reaching significance on MedQA (*r* = 0.079, *p <* 0.01), MedExpQA (*r* = 0.081, *p <* 0.05), Step 1 (*r* = 0.086, *p <* 0.05), and Step 3 (*r* = 0.111, *p <* 0.01), though with smaller effect sizes. In contrast, baseline metrics are largely misaligned with scaling: BLEU-1 (*r* = −0.192, *p <* 0.01), ROUGE-1 (*r* = −0.216, *p <* 0.01), ROUGE-L (*r* = −0.152, *p <* 0.01), METEOR (*r* = −0.089, *p <* 0.01), and BERTScore-F1 (*r* = −0.333, *p <* 0.01) all exhibit strong negative correlations on MedQA, with similar negative patterns across other benchmarks (e.g., BERTScore-F1, *r* = −0.425, *p <* 0.01 on Step 1). BLEU-4 shows only weak and inconsistent positives, i.e., 0.072 on MedQA and 0.091 on MedExpQA, while ROUGE-2 remains near zero throughout. These results confirm that entity-centric coverage metrics, especially EntQA_b_, are the only measures that consistently track systematic quality improvements as model scale increases, whereas conventional overlap-based metrics fail to do so.

**Table 3:** Metric alignment with model scale: Spearman and Kendall correlation analysis across medical benchmarks. This table presents Spearman and Kendall correlation coefficients assessing the relationship between metric scores and model scale on datasets including MedQA, MedExpQA, USMLE Step 1, USMLE Step 2, and USMLE Step 3. Metrics evaluated include entity-based coverage (EntQA_b_ and EntQA_q_), lexical overlap measures (BLEU-1, BLEU-4, ROUGE-1, ROUGE-2, ROUGE-L, METEOR), and semantic similarity (BERTScore-F1). Significance levels are indicated by * (*p <* 0.05) and ** (*p <* 0.01), revealing generally positive correlations for entity-centric metrics and negative associations for most traditional *n*-gram based metrics, suggesting stronger alignment of entity-based evaluations with increasing model scale.

| Metric | MedQA |  | MedExpQA |  | USMLE_Step_1 |  | USMLE_Step_2 |  | USMLE_Step_3 |  |
| --- | --- | --- | --- | --- | --- | --- | --- | --- | --- | --- |
|  | Spearman | Kendall | Spearman | Kendall | Spearman | Kendall | Spearman | Kendall | Spearman | Kendall |
| EntQA<sub>b</sub> | 0.188** | 0.137** | 0.181** | 0.132** | 0.123** | 0.089** | 0.171** | 0.127** | 0.252** | 0.184* |
| EntQA<sub>q</sub> | 0.079** | 0.104** | 0.081* | 0.064* | 0.086* | 0.066* | 0.018 | 0.015 | 0.111** | 0.086** |
| BLEU-1 | -0.192** | -0.138** | -0.106** | -0.076** | -0.331** | -0.241** | -0.219** | -0.158** | -0.177** | -0.128** |
| BLEU-4 | 0.072** | 0.054** | 0.091** | 0.073** | -0.031 | -0.027 | -0.042 | -0.033 | 0.042 | 0.026 |
| ROUGE-1 | -0.216** | -0.156* | -0.113* | -0.082* | -0.359* | -0.261* | -0.244** | -0.176* | -0.213 | -0.154 |
| ROUGE-2 | -0.003 | -0.003 | -0.002 | -0.003* | -0.152** | -0.111* | -0.137** | -0.1* | -0.024 | -0.02 |
| ROUGE-L | -0.152** | -0.109** | -0.088** | -0.064** | -0.306** | -0.224** | -0.207** | -0.149** | -0.117** | -0.084** |
| METEOR | -0.089** | -0.064** | -0.06 | -0.043 | -0.218** | -0.158** | -0.192** | -0.138** | -0.09** | -0.065** |
| BERTScore-F1 | -0.333** | -0.241* | -0.231** | -0.166** | -0.425** | -0.312** | -0.309** | -0.223** | -0.303** | -0.221** |

## 4 Discussion

In this study, we introduced EntQA, a novel entity-centric metric designed to evaluate the clinical alignment and informativeness of LLM-generated responses in medical question-answering (QA) tasks. By leveraging biomedical named entity recognition (NER), semantic embedding-based matching, and coverage score computation, EntQA quantifies how well key entities from patient backgrounds and diagnostic questions are preserved in model outputs, addressing gaps in traditional metrics like accuracy, BLEU, ROUGE, METEOR, and BERTScore. Through experiments on five benchmarks using Qwen 2.5 Instruct models of varying scales, we demonstrated that EntQA exhibits strong positive correlations with model accuracy at both group (Spearman up to 0.9286) and per-case levels (point-biserial up to 0.0926), as well as with model scaling (Spearman up to 0.252). In contrast, baseline metrics often showed negative or inconsistent alignments, underscoring EntQA’s superiority in capturing clinically relevant response quality.

EntQA complements and advances existing evaluation frameworks in medical AI, such as those focused on binary classification metrics like precision, recall and AUC-ROC, which are prevalent in anomaly detection and diagnostic tasks. While these metrics excel in discriminative models, they fall short for generative LLMs, where n-gram-based baselines like BLEU and ROUGE often exhibit negative correlations with accuracy due to their sensitivity to surface-level variations rather than clinical grounding. In contrast, EntQA’s entity-centric approach mitigates common pitfalls, such as overlooking contextual omissions or hallucinations, aligning more closely with trustworthiness metrics proposed for healthcare conversations that emphasize domain-specific fidelity and user-centric evaluation.

The implications of EntQA extend beyond mere benchmarking, offering a pathway to more trustworthy AI-assisted diagnostics in healthcare. By providing interpretable insights into entity omissions (low recall) and hallucinations (low precision), EntQA enables clinicians, researchers and developers to pin-point deficiencies in LLM reasoning, such as overlooking patient-specific details like symptoms or medical history, which could otherwise lead to diagnostic errors. This metric’s focus on topical consistency and grounding in input context supports the development of LLMs that prioritize patient-centered explanations over generic responses, potentially enhancing their utility in real-world clinical workflows, such as decision support systems or educational tools. Furthermore, its extensions to fact-checking and model improvement through entity-conditioned question regeneration, could facilitate iterative fine-tuning of LLMs, promoting safer deployment in high-stakes environments.

Despite these strengths, EntQA has some limitations that warrant consideration. First, its reliance on a biomedical NER model introduces potential biases or errors in entity extraction, particularly for rare or contextually ambiguous medical terms, which could affect coverage scores. The semantic matching thresholds (e.g., cosine similarity *>* 0.7 and lexical overlap *>* 30%) may require dataset-specific tuning to optimize performance, and the metric currently focuses primarily on entity retention without fully capturing higher-level aspects like logical coherence or causal reasoning in responses. Additionally, our evaluation was conducted on a specific family of models (i.e., Qwen 2.5) and English-centric benchmarks, limiting generalizability to multilingual settings, other LLM architectures, or non-QA clinical tasks such as summarization or dialogue. Future work could integrate EntQA into standardized checklists for AI studies, ensuring reproducibility through open-source code and diverse datasets. Additionally, extending EntQA to multimodal tasks (e.g., incorporating imaging or lab data) or combining it with multi-dimensional frameworks, would enhance its applicability to real-world healthcare AI systems.

In conclusion, EntQA represents a step forward in evaluating LLMs for medical QA, outperforming traditional metrics in aligning with accuracy, model scaling, and clinical relevance. By emphasizing entity-level fidelity, it paves the way for more reliable and interpretable AI systems in healthcare, ultimately supporting safer and more effective integration of LLMs into clinical practice.

## Data Availability

All data produced in the present study are available upon reasonable request to the authors.

## Acknowledgments

This project was supported by grants from the National Institute on Aging’s Artificial Intelligence and Technology Collaboratories (P30-AG073105), and the National Institutes of Health (R01-NS142076, R01-HL159620, R01-AG062109, and R01-AG083735).

